# National Increase but Pronounced Regional Disparities in Naloxone Prescribing to United States Medicaid and Medicare Patients

**DOI:** 10.1101/2023.05.17.23290119

**Authors:** Christopher D. Manko, Mohamed S. Ahmed, Lavinia H. Harrison, Sree Kodavatiganti, Noelia Lugo, Jason O. Konadu, Farrin Khan, Carrie A. Massari, Tenisha K. Sealey, Maame Efua Addison, Celine N. Mbah, Kenneth L. McCall, Joseph B. Fraiman, Brian J. Piper

## Abstract

**Background:** Opioid overdoses in the US have increased to unprecedented levels. Administration of the opioid antagonist naloxone can prevent overdoses. This study was conducted to reveal the pharmacoepidemiologic patterns in naloxone prescribing to Medicaid patients from 2018-2021 as well as Medicare in 2019.

**Methods:** The Medicaid State Drug Utilization Data File was utilized to extract information on number of prescriptions and amount prescribed of naloxone at a national and state level. States with naloxone prescription rates differing from the mean by ≥ 1.96 standard deviations were considered statistically significant. The Medicare Provider Utilization and Payment was also utilized to analyze prescription data from 2019.

**Results:** The number of generic naloxone prescriptions per 100,000 Medicaid enrollees decreased 5.15% whereas brand naloxone prescriptions increased 245.00% from 2018-2021. There was a 33.14-fold difference in prescriptions between the highest (New Mexico = 1809.55) and lowest (South Dakota = 54.61) states in 2019. Medicare saw a 30.32-fold difference in prescriptions between the highest (New Mexico) and lowest states (also South Dakota) after correcting per 100,000 enrollees.

**Conclusions:** This pronounced increase in the number of naloxone prescriptions to Medicaid patients from 2018-2021 indicates a national response to this widespread public health emergency. Further research into the origins of the pronounced state-level disparities is warranted.

## Introduction

Opioid overdoses have become a national epidemic in the United States (US). The US exceeded 75,000 opioid-overdose deaths from April 2020 to April 2021 (Centers for Disease Control and Prevention, 2021, November 17). The COVID-19 pandemic appears to have further exacerbated overdoses. A report from Kentucky showed a 71% increase of emergency medical services opioid overdose visits that led to a refusal for transport to the hospital between January and April of 2020 (Slavova, Rock, Bush, Quesinberry, & Walsh, 2020). The cost for opioid addiction and death in the US totaled over one-trillion dollars in 2017 alone (Florence, Luo, & Rice, 2021). Clearly this is a major national issue that needs to be addressed.

The opioid antagonist naloxone can be both easy to administer and immediate in its life-saving effects (Davis & Carr, 2020). Naloxone comes in multiple formulations including injection and nasal spray (Kerensky & Walley, 2017). It is ineffective against other drugs that can cause sedation such as benzodiazepines, alcohol, or non-opioid analgesics (Kerensky & Walley, 2017). However, naloxone is typically benign and does not cause adverse effects when administered to a person who is not overdosing on an opioid (Kerensky & Walley, 2017). Naloxone was first approved by the U.S Food and Drug Administration (FDA) in 1971 (Strang et al., 2019). Since then, it has been formulated with auto-injectors and nasal spray.

Many states have implemented legal mandates to co-prescribe naloxone to individuals at increased risk of overdose (Green, Davis, Xuan, Walley, & Bratberg, 2020). A national evaluation of the number of prescriptions prescribed for naloxone from 2011 to 2017 in IQVIA’s national prescription audit, which contains 90% of all retail pharmacies prescription data (Minji Sohn, Talbert, Huang, Lofwall, & Freeman, 2019) determined that states with naloxone co-prescription laws, had an approximately 7.75-fold higher dispensing rate of naloxone compared to states that did not have the requirement. Nationally, naloxone dispensing to commercially insured patients increased by 13-fold from 7,229 prescriptions in 2015 to 99,917 prescriptions in 2018 (Dunphy, Zhang, Guy, & Jones, 2021).

Due to the continued escalation and potential preventability of opioid overdoses, this study investigated how naloxone prescribing patterns have changed from 2018-2021 among US Medicaid patients. A secondary objective was to characterize any state level disparities among Medicaid and Medicare patients.

## Methods

### Procedures

Prescription data from the State Drug Utilization Data was accessed to review naloxone prescriptions from 2018-2021 (*State Drug Utilization Data*). Enrollee data was also collected from the Centers for Medicare and Medicaid Services (*Medicaid Enrollment Data Collected Through MBES*). The year 2021 was selected as the most recent available at the time of analysis (March 16, 2023 – April 13, 2023). For simplicity, generic naloxone is subsequently designated as Naloxone_G_ and brand name formulations as Naloxone_B_. Cost of Naloxone_B_ and Naloxone_G_ per prescription over time was also obtained.

The Medicare Provider Utilization and Payment database was used to extract drug prescription claims as well as location for the year 2019 (*Medicare Part D Prescribers*). This Medicare database extracts information from finalized prescription drug claims, which excludes any claims that have not been resolved or need adjustments that have not been completed. In addition, the database suppresses any provider and claim data for less than or equal to eleven total claims for that year and represented by a blank value. Enrollees for that year were also collected from KFF’s dataset (*Total Number of Medicare Beneficiaries by Type of Coverage*). Data was collected from March 16, 2023 – April 26, 2023. This study was approved as exempt by the Geisinger IRB.

### Data Analysis

The prescriptions of naloxone per state were corrected for Medicaid enrollment which was pulled from the first month of each quarter from the database (*Medicaid Enrollment Data Collected Through MBES*). National distribution was plotted versus time using GraphPad Prism. A heatmap and waterfall graph were created to visualize disparities in distribution. States outside of a 95% confidence interval (mean ± 1.96*SD) were considered elevated although states outside ±1.5 SD were also noted. Medicare’s data was also corrected for the number of Part D enrollees.

## Results

### Medicaid

Figure 1 shows an overall increase in quarterly prescriptions after correcting for Medicaid enrollees. When comparing corrected national prescriptions of Naloxone_B_ and Naloxone_G_, a distinct difference was noted. Q1 of 2018 (93.54) to Q4 2021 (322.71) for Naloxone_B_ showed a large increase (+245.00%). For Naloxone_G_ it decreased from 14.95 to 14.18 (−5.15%). When combined, Q1 of 2018 was calculated as 108.49 and Q4 of 2021 was 336.89. Thus, a 3.11-fold increase was seen in those four years.

**Figure 1.**
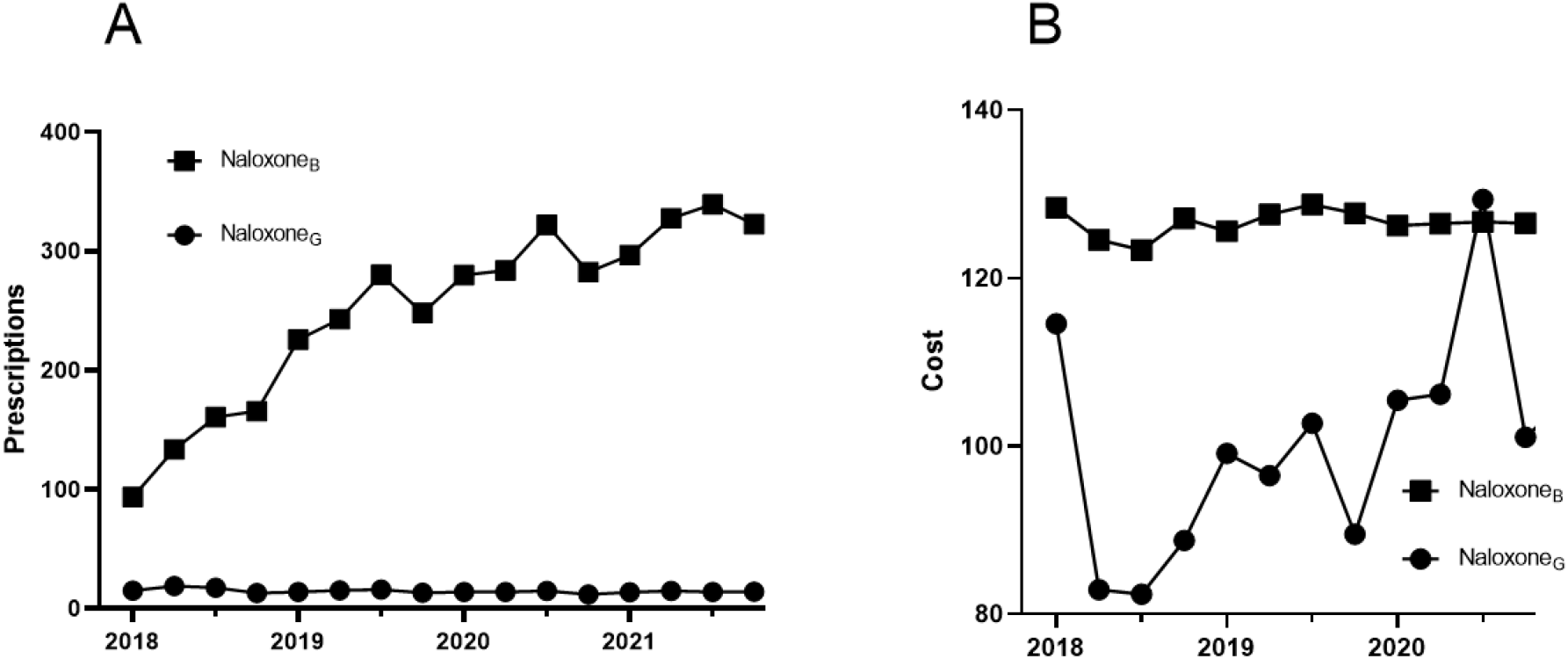
(A) Total naloxone prescriptions in Medicaid by formulation per quarter corrected for number of enrollees. (B) Cost per prescription ($USD) of generic (Naloxone_G)_ and brand (Naloxone_B)_ formulations for 2018 to 2021.

Cost of prescription was also assessed. Naloxone_B_ showed a slight (−1.58%) decrease while Naloxone_G_ decreased by 14.63% from 2018 to 2021. The cost per prescription of Naloxone_G_ decreased from $114.59 in the beginning of 2018 to $97.82 at the end of 2021. Naloxone_B_ slightly decreased from $128.41 to $126.38.

Examination of prescription rates by state in 2019 were also completed (Figure 2). The top five highest prescribing states were New Mexico, Rhode Island, Maryland, Pennsylvania, and Delaware. Z-scores were calculated, and Delaware was 1.5 standard deviations above the mean, whereas the other 4 states were 1.96 standard deviations above the mean. There was a 33.14-fold greater prescribing rate in New Mexico (1,809.55) prescriptions per 100k enrollees) relative to South Dakota (54.61).

**Figure 2.**
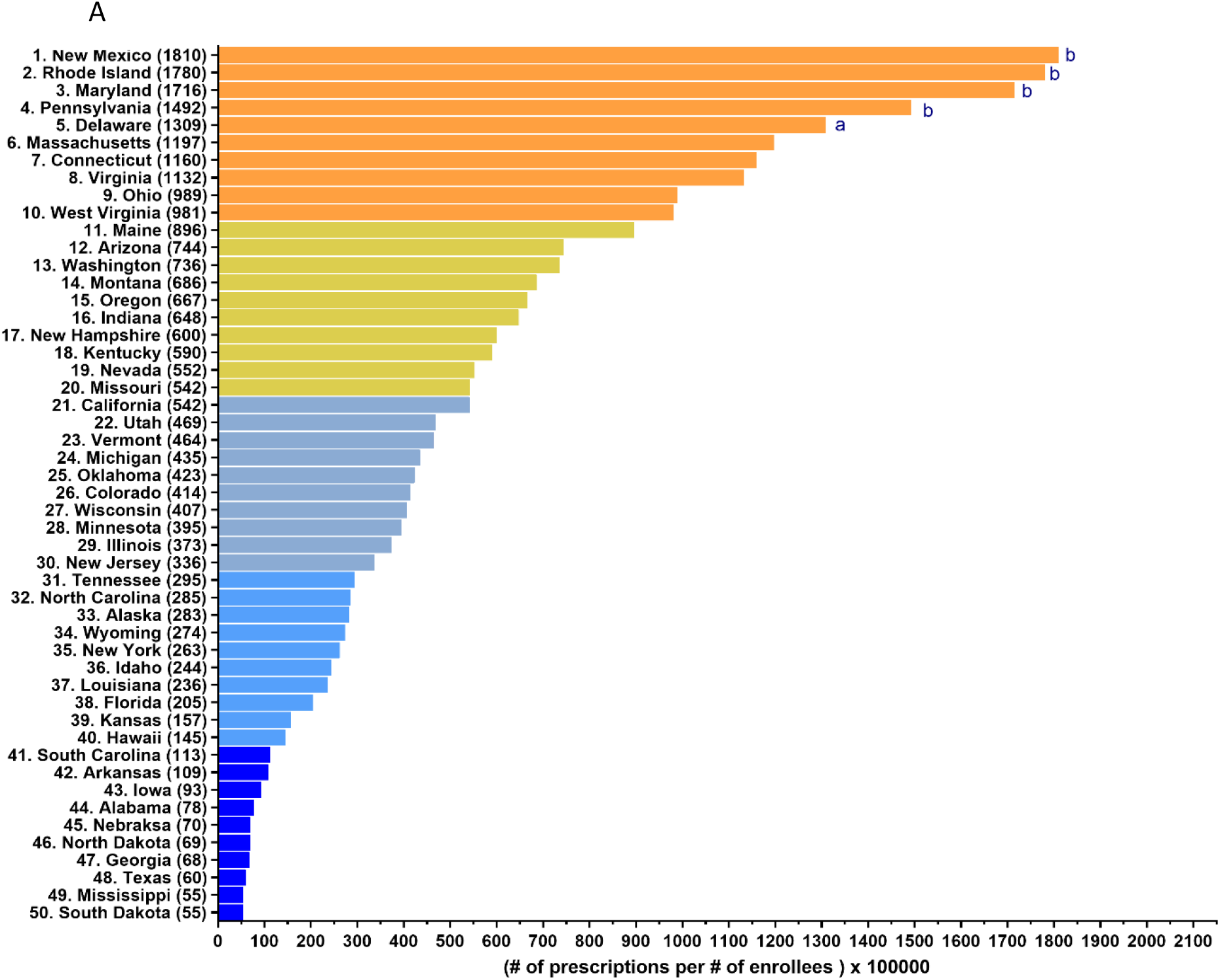
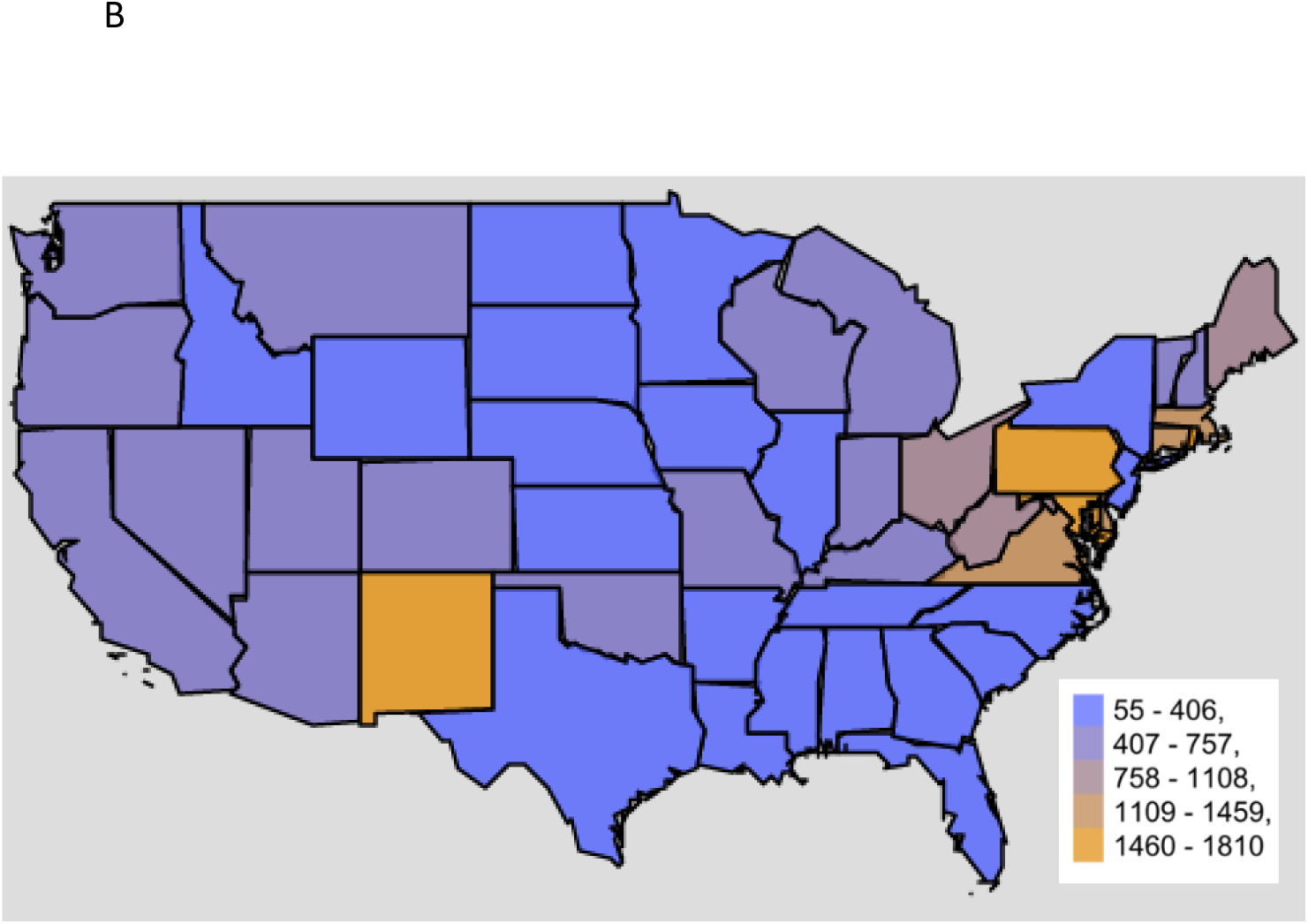
Naloxone prescriptions per 100,000 Medicaid enrollees by state in 2019 as a waterfall (A) and heatmap (B). ^a^≥1.5 standard deviations above the mean, and ^b^≥1.96 standard deviations above the mean (552.84).

### Medicare

When calculating through Z-score in the Medicare data for 2019, New Mexico and California were ≥ 1.96 standard deviations above the mean (Figure 3). After adjusting total claims per 100,000 beneficiaries per state, New Mexico, California, Tennessee, Rhode Island, and Arizona were the top prescription states. There was a 30.32-fold difference between the highest (New Mexico) and the lowest (South Dakota) states.

**Figure 3.**
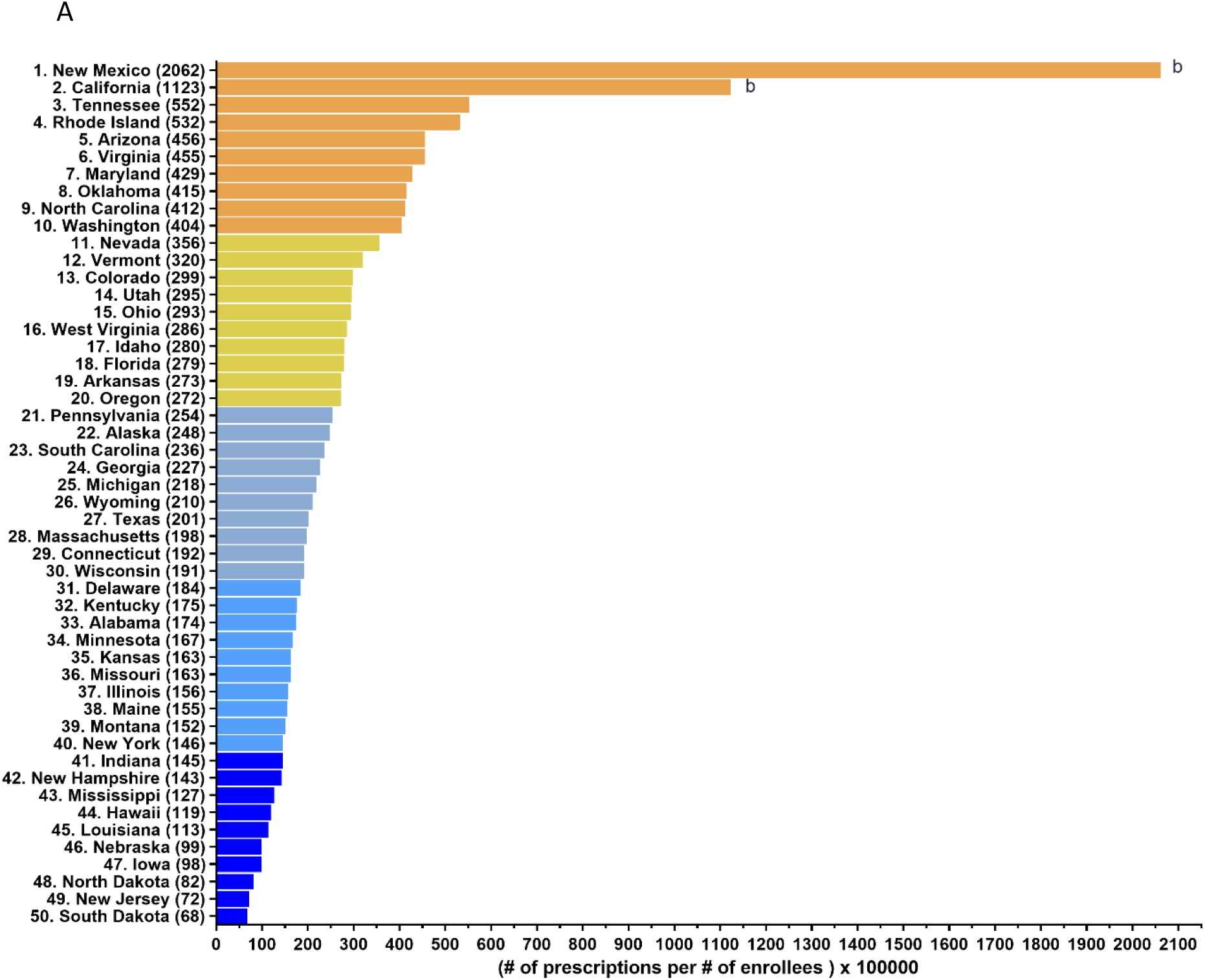
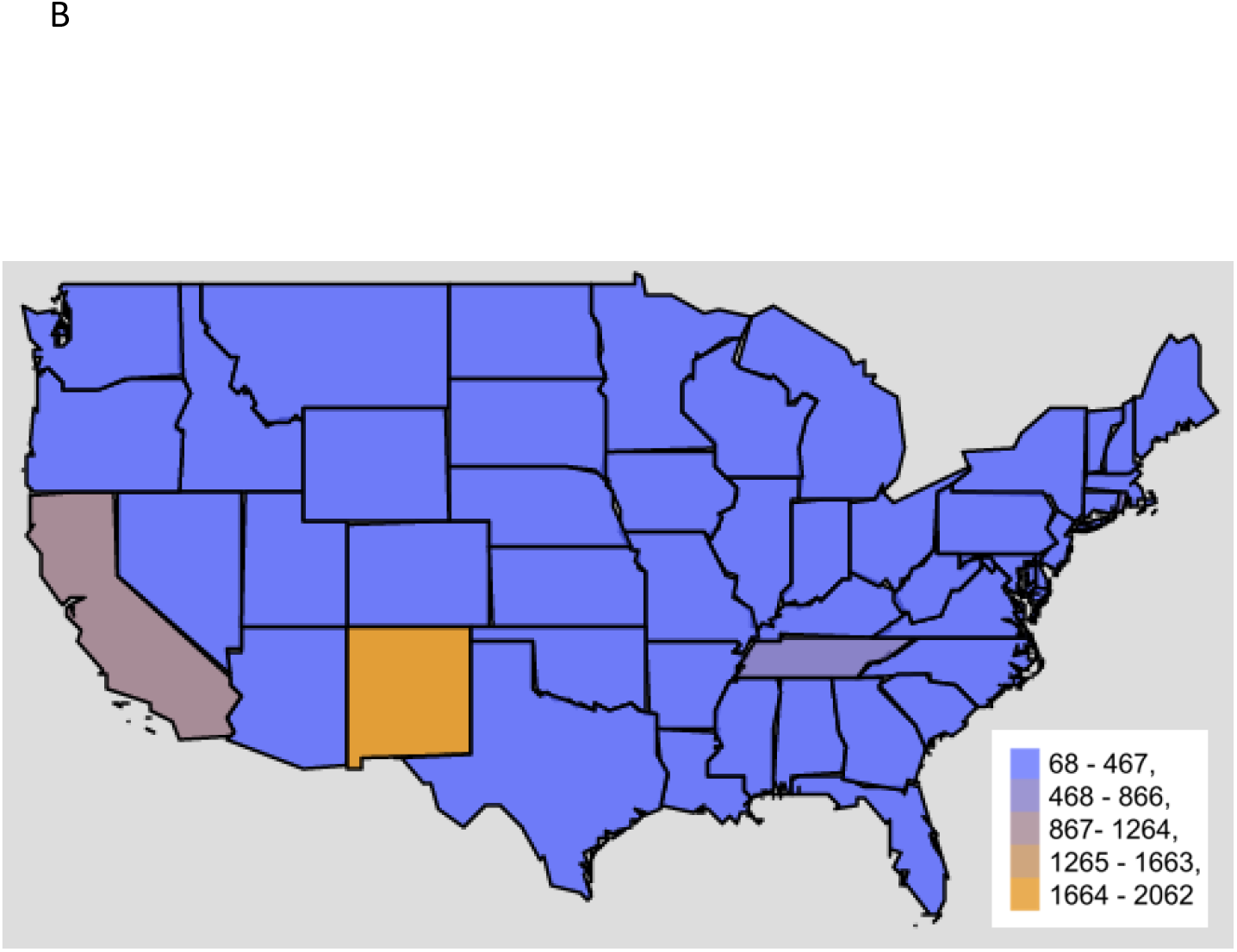
Naloxone prescriptions per 100,000 Medicare enrollees per state in 2019 as a waterfall (A) and heatmap (B). ^a^≥1.5 standard deviations above the mean, and ^b^≥1.96 standard deviations above the mean (293.38).

Finally, a scatterplot was created to examine the association of the Medicaid and Medicare prescribing (Figure 4). The r value was calculated as 0.472, p<0.001. When NM and CA were removed, the correlation was found to be r(46) = 0.432, p<0.01.

**Figure 4.**
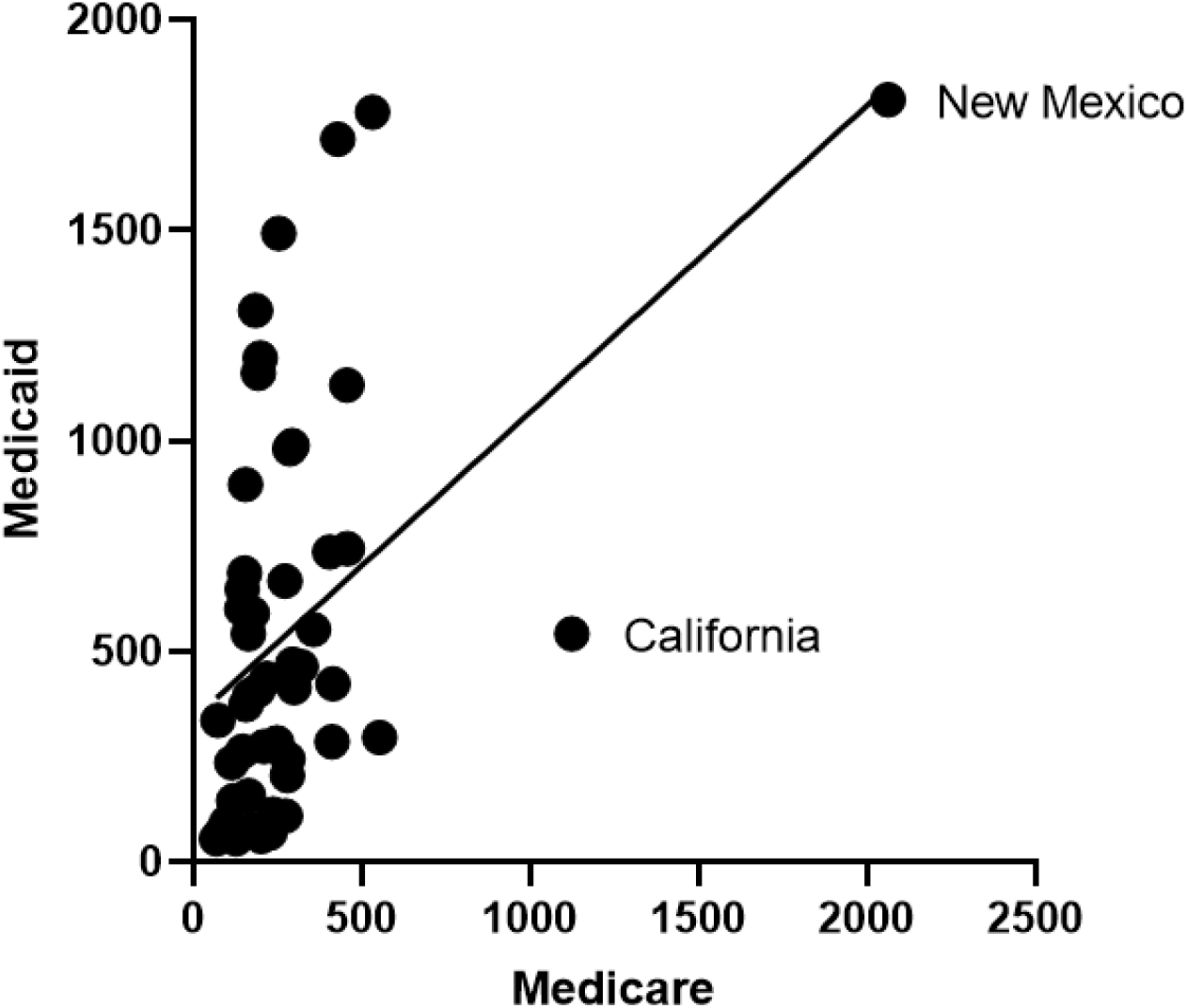
Medicare and Medicaid naloxone claims per state correcting for the number of enrollees (r(48) = 0.472, p<0.001).

## Discussion

Overall, there was a substantial increase in naloxone prescriptions. This 210.53% elevation in Medicaid should come as no surprise for a few reasons. Past work has similarly described increases in naloxone prescriptions within Medicaid during an earlier (2013-2017) period (Roberts, 2021). The increase in prescription may be due to Medicaid expansion and the updates in standing orders/laws individual states issue. Medicaid expansion accounted for an 8.3% increase in total naloxone prescriptions (Frank & Fry, 2019). Further review of the literature has only continued to solidify the weight of Medicaid’s influence over naloxone access (M. Sohn et al., 2020). Individuals insured by Medicaid may face financial and housing insecurity (Tilhou, Dague, Saloner, Beemon, & Burns, 2022). As such, coverage of naloxone may be a major factor for accessibility. State interventions such as standing orders show conflicting evidence over their efficacy in increasing accessibility to naloxone (Gertner, Domino, & Davis, 2018; Xu, Davis, Cruz, & Lurie, 2018). Internationally, we see increased naloxone accessibility with implementation of programs such as in Ontario (Choremis et al., 2019). However, while there are studies in favor of state intervention, other studies have argued otherwise. For example, in North Carolina, one report showed evidence that even with the implementation of a standing order, only three-fifths of retail pharmacies carried naloxone (Egan, Foster, Knudsen, & Lee, 2020). Another report from Philadelphia revealed that communities that had higher rates of opioid overdose were less likely to have pharmacy accessibility to naloxone (Guadamuz, Alexander, Chaudhri, Trotzky-Sirr, & Qato, 2019).

This study also showed the price of generic naloxone increased. Multiple variables can account for this rise. Increased demand, drug shortages, and limited competition may all influence this increase (Rosenberg, Chai, Mehta, & Schick, 2018).

Finally, when looking at state-by-state naloxone prescription rates, disparities were clearly seen throughout the US. There was a 33.14-fold difference between the highest and lowest states when correcting for enrollees in 2019 for Medicaid. While not as large, Medicare data still showed a 30.32-fold difference. Literature investigating naloxone patterns in the past have provided many arguments as to why. Some major themes highlighted, which may be present here, include financial barriers, standing orders/state intervention, and the stigma/guidance providers are exposed to. As mentioned previously, the population insured by Medicaid may face a variety of issues such as financial and housing insecurity (Frank & Fry, 2019). Thus, the actual coverage and expansion Medicaid provides carries appreciable influence. If we assume each state has their own funded programs to assist community members, along with differing levels of efficacy in education on such resources, we can already begin to identify discrepancies between states. One study provided evidence that even within a specific area, community, pharmacy, and prescription-based access to naloxone have differing levels of effectiveness at increasing accessibility (Irvine et al., 2022). Finally, stigma and guidance can have a profound effect on naloxone accessibility. For example, pharmacies in Texas not only differ on naloxone access, but also on reasons for why they are not or will not be available (Hill et al., 2021). For the consumer, this can make accessing naloxone very challenging, and reasons for such barriers may be rooted in stigma. One pharmacist stated, “being that kind of medication, we are going to pass.” (Hill et al., 2021). States may provide legal access for consumers, but if stigma runs rampant, people who need naloxone will face major challenges getting it. Influence can have an opposite impact too. For example, one report noted how a recommendation by the American Society of Anesthesiologists led to an increase in prescriptions of naloxone (Xu et al., 2018). Hence, state-by-state levels of stigma contribute to the differing levels of naloxone prescription, and this is further affected by different professional association recommendations and guidelines for health care providers.

Our distinction between brand and generic versions of naloxone showed major differences in both prescriptions and cost. Given the population of Medicaid patients and the financial struggles they may face, financial barriers imposed by pharmaceutical companies can bar patients from accessing this life saving drug. Of all opioid overdoses, 30% of patients are uninsured, thus making up an important subpopulation where finances can make a large impact (Peet, Powell, & Pacula, 2022). In comparison, for the buprenorphine market it was found that multiple factors inhibited the release of generic sublingual buprenorphine (Barenie, Sinha, & Kesselheim, 2021). Additionally, when examining insured patients specifically, the cost of buprenorphine decreased overtime (Roberts, Saloner, & Dusetzina, 2018). This interesting pattern follows naloxone, which, for insured patients decreased by 26.15% (Peet et al., 2022). However, for the uninsured patient desiring naloxone, they instead saw a staggering 506.33% increase in cost (Peet et al., 2022). As such, future work should be done to encourage generic formulations of products that can combat the opioid epidemic and should also work to make prices more affordable to uninsured patients.

Some caveats and future directions are noteworthy. This study characterized the increases and pronounced state-level variation to Medicaid patients. As naloxone is prescribed for those at risk of opioid overdose and patients with an opioid use disorder, the denominator in the analyses was the number of patients in each program (Medicaid or Medicare). Further research should further examine the sources of naloxone (e.g. primary care vs specialists) and specific patient populations (those with a history of emergency room visits for overdoses). As there are regional differences (East vs West) in overdoses and fentanyl seizures (Zoorob, 2019), further investigations of how naloxone prescribing has impacted opioid mortality, before and during COVID-19 pandemic, is needed.

## Conclusion

While naloxone prescriptions to Medicaid patients have increased, more work should clearly be done to target this crisis including identifying the origins for the 33.14-fold disparities in prescribing between states and maximizing availability of evidence-based treatments to resource limited populations. Education, preparedness to respond to overdose, and increased co-prescription are different avenues worth further exploration.

## Data Availability

All data produced in the present study are available upon reasonable request to the authors.

https://www.medicaid.gov/medicaid/prescription-drugs/state-drug-utilization-data/index.html

https://www.medicaid.gov/medicaid/national-medicaid-chip-program-information/medicaid-chip-enrollment-data/medicaid-enrollment-data-collected-through-mbes/index.html

https://data.cms.gov/provider-summary-by-type-of-service/medicare-part-d-prescribers

https://www.kff.org/medicare/state-indicator/total-medicare-beneficiaries/?currentTimeframe=0&sortModel=%7B%22colId%22:%22Location%22,%22sort%22:%22asc%22%7D

## Acknowledgements

This research was generously supported by the Health Resources and Services Administration (D34HP31025).

## Disclosures

Brian J. Piper was (2019-21) part of an osteoarthritis research team supported by Pfizer and Eli Lilly. The other authors have no disclosures.

## Notes

### Author Declarations

The study used enrollee and prescription data that is publicly available since the start of data collection.

